# Impact of Glucose Trajectories on Outcomes After Intracerebral Hemorrhage: The ATACH-2 trial

**DOI:** 10.64898/2026.03.12.26348296

**Authors:** Mahmoud Fayed, Neha Saini, Sebastian Edwards, Cheng Zeng, Leo Duan, Amita Singh, Anna Khanna, Christina A. Wilson, Adnan I. Qureshi, Teng J. Peng

**Affiliations:** University of Florida, College of Medicine, Department of Neurology, Gainesville, FL, 32601; University of Florida, Department of Biostatistics, Gainesville, FL, 32601; Zeenat Qureshi Stroke Institute, Columbia, MO 65211; Department of Neurology, University of Missouri, Columbia, MO, 65212

**Author notes:** **Correspondence:** Teng J. Peng, MD, 1329 SW 16^th^, Street, Suite 1160, Gainesville, FL, 32608.

**Keywords:** [intracerebral hemorrhage], [stroke], [glucose]

## Abstract

**Background:** Hyperglycemia after intracerebral hemorrhage (ICH) may be associated with worse outcomes. In this study, we evaluated the association of early post-ICH glucose trajectories and clinical outcomes.

**Methods:** We performed a secondary analysis of the ATACH-2 trial dataset. Hyperglycemia was defined as a blood glucose of ≥140 mg/dl. Glucose levels at 0h, 24h, 48h, and 72h were analyzed using a linear mixed effects model, with fixed effects for time and random intercept/slopes. Patient-specific estimates were used to predict glucose values at 0h and 72h, informed by all four timepoints, to classify patients into the following glycemic trajectory groups: (1) early hyperglycemia, (2) late hyperglycemia, (3) persistent hyperglycemia, and (4) persistent normoglycemia. Outcomes were compared using univariate analysis and log-rank test survival analysis. Good outcomes were defined as a modified Rankin Score of 0 to 2. The association between glycemic trajectories and functional outcomes was tested using logistic regression models adjusted for patient demographics and clinical variables.

**Results:** Of 1000 patients (median age 62 [IQR 52–71]; 38% female) in the study, 81 (8.1%) had early hyperglycemia, 59 (5.9%) late hyperglycemia, 225 (22.5%) persistent hyperglycemia, and 635 (63.5%) persistent normoglycemia. On univariate analysis, 45.8% of patients with persistent normoglycemia had favorable 90-day functional outcomes compared to 30.9% in early, 30.5% in late, and 32.0% in persistent hyperglycemia patients (p<0.001). The late hyperglycemia patients had the highest rate of hematoma expansion (35.3%, p=0.029) and the lowest Kaplan Meier-estimated survival (86%, p=0.015). In adjusted multivariable regression models, early hyperglycemia was significantly associated with a poor functional outcome (OR 2.27, 95% CI 1.10-4.68, p=0.026).

**Conclusion:** Early hyperglycemia was associated with worse functional outcomes, while late and persistent hyperglycemia were associated with worse survival rates. These findings suggest that glycemic trajectories may affect or predict prognosis. This highlights the importance of continuous glucose monitoring and glycemic control strategies after ICH.

## Introduction

Intracerebral hemorrhage accounts for approximately 15% of all strokes and is associated with a 30-day mortality rate exceeding 30%.^1^ Standard acute management centers on blood pressure control, reversal of coagulopathy, treatment of seizures, and correction of metabolic derangements.^2^ However, the optimal management of metabolic derangements remains less defined. While admission hyperglycemia has been associated with the severity of ICH and is a well-established predictor of poor functional outcomes in both diabetic and non-diabetic patients,^3,4,5^ this approach fails to capture the dynamic nature of post-ICH metabolic stress.^6^

Recent evidence suggests that delayed hyperglycemia at 24 and 72 hours and high glycemic variability over the first seven days of hospital admission are more robustly associated with 90-day functional impairment and 30-day neurological deterioration.^7,8^ Therefore, measurement of glycemic change within the first few days after admission could offer better prognostic factors than a single glucose test on admission.^9,10,11^ Current American Heart Association (AHA) guidelines recommend general glucose monitoring and avoidance of hypoglycemia or hyperglycemia without providing specific glycemic targets or management protocols.^2^ There remains a lack of literature regarding the specific impact of glycemic changes on ICH-related outcomes. This study aims to examine the association between blood glucose trajectories and ICH-related outcomes.

## Methods

We performed a secondary analysis of the ATACH-2 trial. Data was obtained from the NINDS biorepository. This study was exempt from the UF IRB ET00049461. This study was reported in accordance with the STROBE guidelines for observational studies. Statistical analysis was performed using R version 4.5.1 (R Foundation for Statistical Computing, Vienna, Austria, https://www.r-project.org).

### Population and Study Design

The ATACH-2 trial is a prospective, multicenter, randomized, open-label study that aims to determine the safety and superiority of intensive blood pressure management in patients with intracerebral hemorrhage presenting within 4.5 hours of symptom onset. Participants were randomized to intensive blood pressure management with a target of 110-139mmHg or standard blood pressure management with a target of 140-179mmHg. The trial enrolled 1000 patients from May 2011 to September 2015. Inclusion and exclusion criteria have been previously published.^12^

### Outcomes

The primary outcome of this study is to evaluate if early glucose trajectories were associated with worse functional outcome, defined as mRS of 3-6. Secondary outcomes include evaluating if admission hyperglycemia was associated with worse outcomes, perihematomal edema, and hematoma expansion.

### Variable Definitions

In this study, hyperglycemia was defined as a blood glucose level of ≥140 mg/dl.^13^ Glucose was measured at baseline, 24h, 48h, and 72h after hospital arrival. Hematoma expansion was defined as an increase of ≥33% in ICH,^13,14^ and perihematomal edema expansion as an increase of ≥40% in perihematomal edema volume on the 24h computed tomography (CT) scan compared to the baseline CT scan.^14^

### Determination of Glucose Trajectories

To identify patterns in glucose level changes across these time points, we fitted a linear mixed effects model.^15^ This model incorporates both fixed and random effects, allowing us to estimate overall glucose trends across patients while accounting for individual differences. We modeled glucose as a linear function of time, with a random slope and intercept for each patient. After fitting the model, we used the patient-specific estimates to predict glucose values at 0h and 72h, which were informed by all four time points. Based on the predicted values at 0h and 72h, we categorized each patient into one of four glycemic trajectory groups: (1) early hyperglycemia, defined as having a predicted glucose level of 140 mg/dL or higher at 0h and below 140 mg/dL at 72h; (2) late hyperglycemia, defined as having a predicted glucose level below 140 mg/dL at 0h and 140 mg/dL or higher at 72h; (3) persistent hyperglycemia, defined as having predicted glucose levels of 140 mg/dL or higher at both 0h and 72h; and (4) persistent normoglycemia, defined as having predicted glucose levels below 140 mg/dL at both time points.

### Statistical Analyses

Patients were stratified by their glucose trajectory group, and univariate analysis was used to compare patient baseline characteristics, stroke severity, imaging metrics, and functional outcomes (Table 1). Chi-square test was used for categorical variables and Mann-Whitney test for continuous variables due to the non-normal distribution of data. We used numbers (percentages) to express categorical variables and median (interquartile range, IQR) for continuous variables. Functional outcomes were measured through modified Rankin Scale (mRS) score at 90 days and were dichotomized into good functional outcomes (mRS score of 0-2) and poor functional outcomes of disability or death (mRS scores of 3-5 and 6, respectively).

**Table 1.** Baseline characteristics of patients across different glucose trajectory groups.

|  |  | Early<br>Hyperglyce<br>mia | Late<br>Hyperglyce<br>mia | Persistent<br>Hyperglyce<br>mia | Persistent<br>Normoglyce<br>mia | p |
| --- | --- | --- | --- | --- | --- | --- |
| n |  | 81 | 59 | 225 | 635 |  |
| Age (median [IQR]) |  | 60.00<br>[51.00,<br>70.00] | 60.00<br>[52.00,<br>72.00] | 63.00<br>[53.00,<br>72.00] | 62.00<br>[52.00,<br>70.00] | 0.559 |
| Sex (%) | Male | 48 (59.3) | 33 (55.9) | 149 (66.2) | 390 (61.4) | 0.396 |
|  | Female | 33 (40.7) | 26 (44.1) | 76 (33.8) | 245 (38.6) |  |
| Race (%) | White | 27 (33.3) | 16 (27.1) | 66 (29.3) | 178 (28.0) | 0.904 |
|  | Black | 11 (13.6) | 7 (11.9) | 29 (12.9) | 86 (13.5) |  |
|  | Asian | 41 (50.6) | 36 (61.0) | 124 (55.1) | 361 (56.9) |  |
|  | Other | 2 (2.5) | 0 (0.0) | 6 (2.7) | 10 (1.6) |  |
| Stroke/TIA (%) |  | 9 (11.1) | 9 (15.3) | 52 (23.3) | 94 (14.9) | **0.015** |
| Atrial Fibrillation (%) |  | 8 (9.9) | 3 (5.1) | 9 (4.0) | 16 (2.6) | **0.009** |
| Hypertension (%) |  | 63 (81.8) | 48 (82.8) | 190 (88.0) | 492 (79.1) | **0.038** |
| Hyperlipidemia (%) |  | 23 (29.9) | 13 (23.6) | 73 (34.6) | 132 (22.1) | **0.003** |
| Diabetes (%) |  | 17 (21.0) | 12 (20.7) | 127 (58.3) | 30 (4.8) | **<0.001** |
| Smoking (%) | Current | 22 (29.3) | 11 (19.6) | 50 (24.5) | 181 (31.3) | **<0.001** |
|  | Former | 15 (20.0) | 5 (8.9) | 57 (27.9) | 98 (16.9) |  |
|  | Never | 38 (50.7) | 40 (71.4) | 97 (47.5) | 300 (51.8) |  |
| Treatment<br>arm (%) | Intensive | 57 (70.4) | 31 (52.5) | 105 (46.7) | 307 (48.3) | **0.002** |
|  | Standard | 24 (29.6) | 28 (47.5) | 120 (53.3) | 328 (51.7) |  |
| NIHSS at baseline<br>(median [IQR]) |  | 11.00 [5.75,<br>16.00] | 13.00 [9.00,<br>19.00] | 12.00 [6.75,<br>16.25] | 10.00 [6.00,<br>15.00] | **0.009** |
| GCS at baseline (median<br>[IQR]) |  | 15.00<br>[13.00,<br>15.00] | 14.00<br>[12.50,<br>15.00] | 15.00<br>[12.00,<br>15.00] | 15.00<br>[14.00,<br>15.00] | **0.01** |
| Hemorrhage<br>location (%) | Basal<br>ganglia | 43 (53.1) | 35 (59.3) | 121 (53.8) | 363 (57.3) | 0.31 |
|  | Thalamus | 31 (38.3) | 13 (22.0) | 76 (33.8) | 200 (31.6) |  |
|  | Lobar | 7 (8.6) | 11 (18.6) | 28 (12.4) | 70 (11.1) |  |
| IVH (%) |  | 27 (33.3) | 15 (25.4) | 78 (34.7) | 165 (26.0) | 0.061 |
| ICH volume at baseline<br>(cm) (median [IQR]) |  | 11.61 [6.10,<br>21.25] | 16.22 [9.67,<br>22.55] | 10.91 [5.39,<br>21.51] | 9.12 [4.59,<br>16.66] | **<0.001** |
| Perihematoma edema<br>volume at baseline (cm)<br>(median [IQR]) |  | 1.36 [0.57,<br>2.66] | 1.34 [0.67,<br>2.53] | 1.45 [0.76,<br>2.84] | 1.38 [0.65,<br>2.42] | 0.405 |
| Hematoma expansion (%) |  | 9 (12.7) | 18 (35.3) | 42 (21.8) | 120 (21.4) | **0.029** |
| IVH expansion (%) |  | 5 (21.7) | 10 (58.8) | 34 (44.2) | 55 (35.9) | 0.067 |
| Perihematomal edema expansion (%) |  | 31 (43.7) | 32 (62.7) | 99 (51.3) | 292 (52.0) | 0.225 |
| mRS at 90 days (%) | 0-2 | 25 (30.9) | 18 (30.5) | 72 (32.0) | 291 (45.8) | **<0.001** |
|  | 3-6 | 56 (69.1) | 41 (69.5) | 153 (68.0) | 344 (54.2) |  |
| Mortality at 90 days (%) |  | 5 (6.2) | 8 (13.6) | 22 (9.8) | 32 (5.0) | **0.014** |
| Hospital LOS (median [IQR]) |  | 14.00 [7.00, 25.25] | 20.00 [12.50, 36.50] | 17.00 [8.00, 29.00] | 14.00 [7.00, 24.00] | **0.001** |

LOS = Length of Stay

We chose mRS score of 0-2 as a cut-off for good functional outcomes because these patients do not need assistance in performing activities of daily living. For survival analysis and logistic regression, we excluded 39 patients who had a missing 90-day mRS score. Based on prior studies, we treated NIHSS score as a continuous variable graded from 0 to 42.^16^

Survival difference between the glycemic trajectories was assessed using log-rank test and plotted on a Kaplan-Meier curve. Time from randomization to death was measured in days, and patients who were alive at 90 days were censored at that time point. We performed a univariable Cox proportional hazards regression analysis to test for the association between glycemic trajectory group and death within 90 days by estimating hazard ratios (HRs) for each glycemic group.

Association of glycemic trajectories with functional outcome was tested using logistic regression models. We first used a univariable model to test for crude association. Then we used a multivariable model to adjust for variables of univariate significance (p<0.1) and/or clinical relevance such as age, sex, race, past medical history of stroke, transient ischemic attack (TIA), diabetes mellitus, hypertension, and smoking, treatment arm, presence of IVH, ICH volume, ICH location, National Institutes of Health Stroke Scale (NIHSS) score at baseline, Glasgow Coma Scale (GCS) score. Persistent normoglycemia was used as the reference group for both the Cox and logistic regression models. We reported odds ratios (ORs) with 95% confidence intervals (CIs) and p-values. A two-sided p<0.05 was considered statistically significant.

### Subgroup Analyses

As a subgroup analysis, we repeated the analysis in patients with and without diabetes to evaluate differences between chronic hyperglycemia and stress-induced hyperglycemia. Since blood pressure treatment could be a confounding factor, we also performed a subgroup analysis of patients in the treatment vs control arm.

## Results

### Univariate analysis

Our cohort had a median age of 62 (IQR 52-71) and was 38% female. Race was predominantly Asian (52.6%), followed by White (28.7%) and Black (13.3%). Of the 1000 patients in our analysis, 81 (8.1%) had early hyperglycemia, 59 (5.9%) late hyperglycemia, 225 (22.5%) persistent hyperglycemia, and 635 (63.5%) persistent normoglycemia (Table 1). On univariate analysis, 45.8% of patients with persistent normoglycemia had favorable 90-day functional outcomes compared to 30.9% in early, 30.5% in late, and 32.0% in persistent hyperglycemia patients (p<0.001) (Figure 1). The late hyperglycemia group had the highest rate of hematoma expansion at 35.3% (p=0.029). Late and persistent hyperglycemia groups had the highest rates of IVH expansion, both at around 24% (p<0.001).

**Figure 1.**
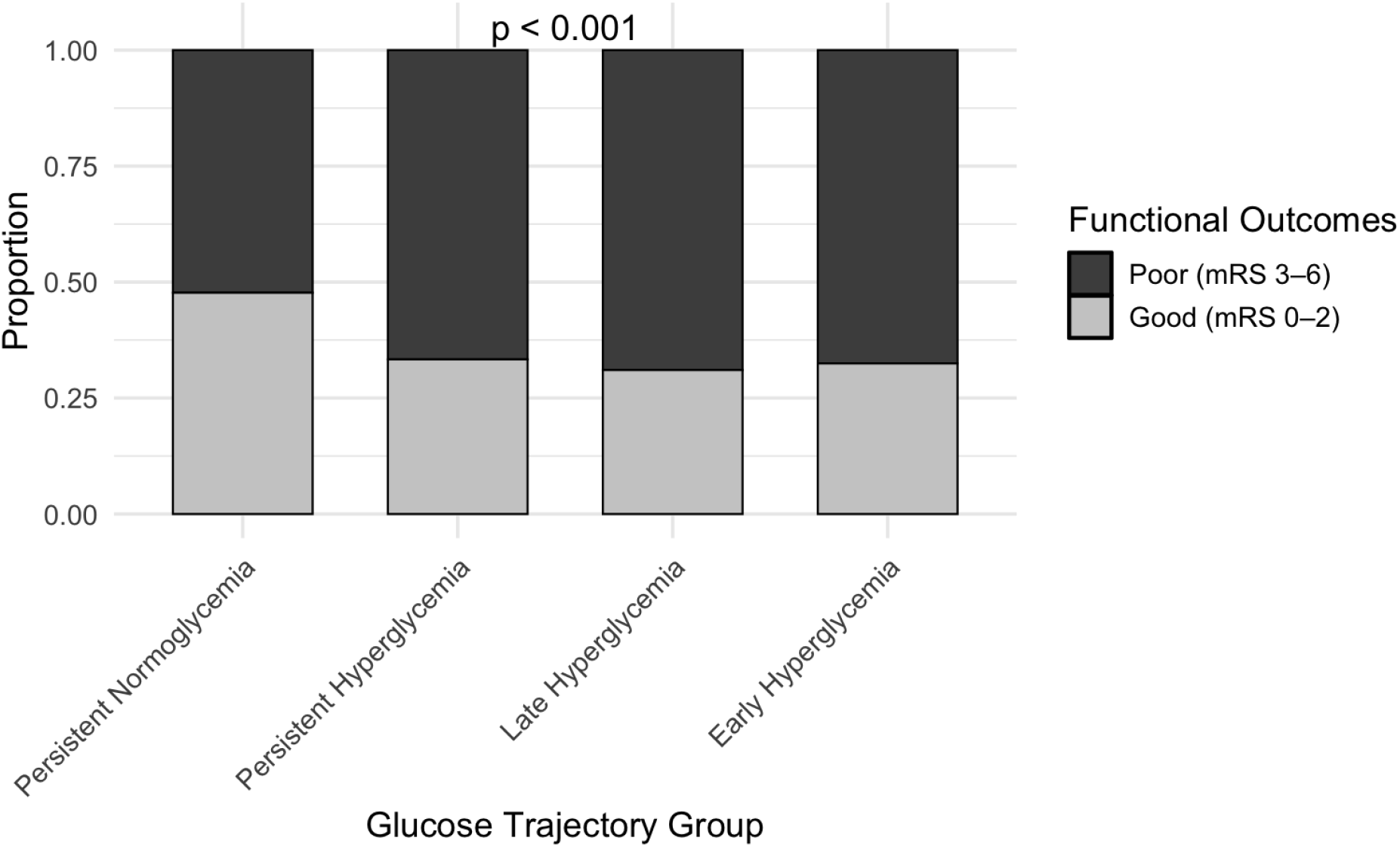
Functional outcomes by glucose trajectory groups. Patients with missing 90-day functional outcomes were excluded

### Survival analysis

Among the 961 patients that had a 90-day follow-up, there was a significant difference in the survival probability between the different glucose trajectory groups (p=0.015). Patients with late hyperglycemia showed the lowest Kaplan-Meier–estimated survival probability (86%), followed by those with persistent hyperglycemia (90%) (Figure 2). In the univariable Cox proportional hazards regression model, the late hyperglycemia group patients and persistent hyperglycemia patients were 2.71 times and 2.00 times more likely to die within 90 days (95% CI 1.25-5.87, p=0.012, and 95% CI 1.16-3.43, p=0.013, respectively).

**Figure 2.**
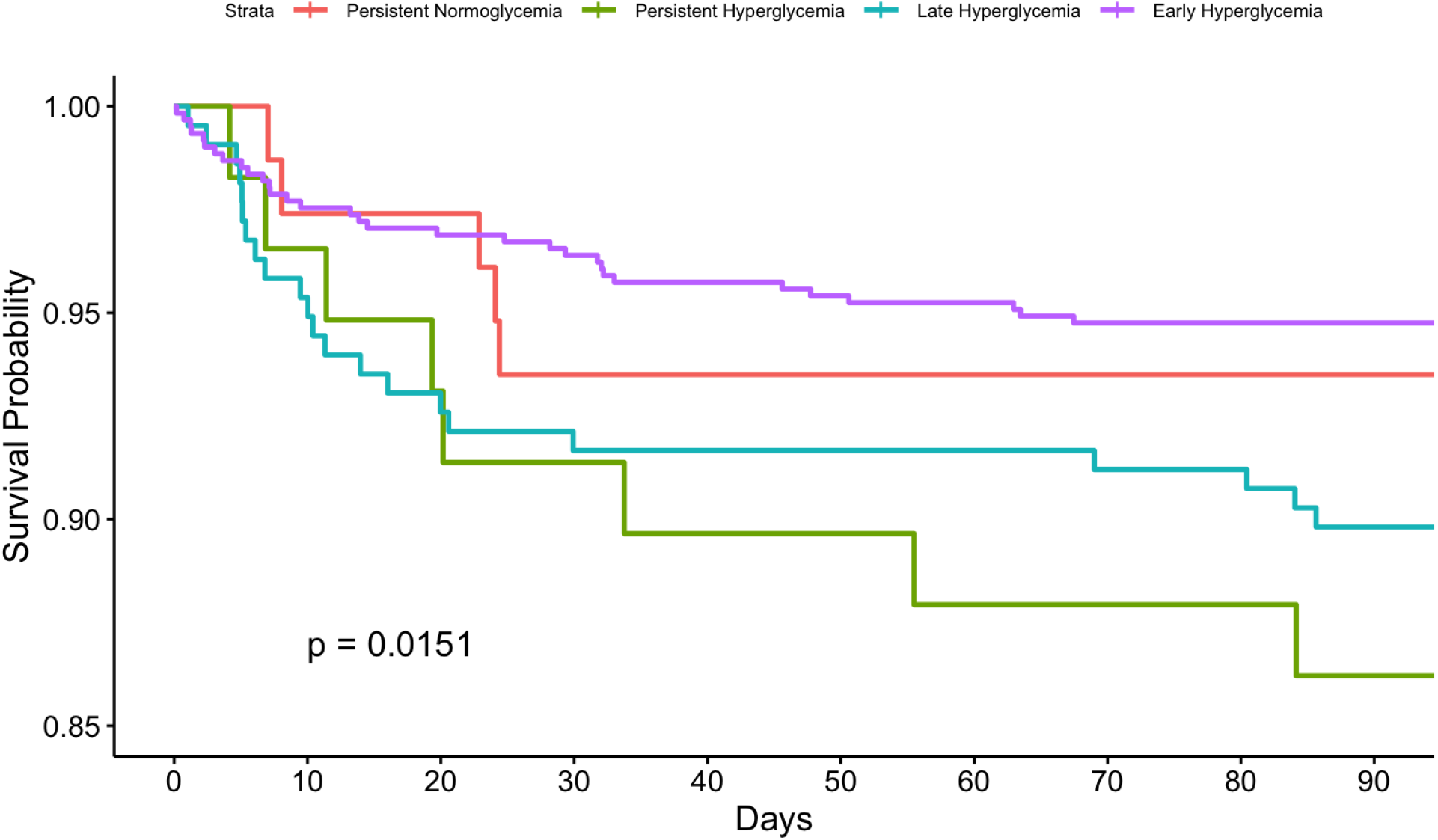
Kaplan–Meier survival curves by glucose trajectory groups.

### Logistic regression

In univariate analysis, early hyperglycemia (OR 1.90, 95% CI 1.15-3.14, p=0.013), late hyperglycemia (OR 2.03, 95% CI 1.14-3.62, p=0.017), and persistent hyperglycemia (OR 1.82, 95% CI 1.32-2.52, p<0.001) were all associated with worse outcomes (Table 2). In the adjusted model, early hyperglycemia remained statistically significantly associated with a poor functional outcome (OR 2.27., 95% CI 1.10-4.68, p=0.026) (Table 3). Other notable variables with statistically significant association with poor functional outcomes included age, female sex, baseline NIHSS, basal ganglia and thalamic hemorrhage, presence of IVH, and past medical history of hypertension. Asian populations were associated with better functional outcomes compared to white populations.

**Table 2.** Unadjusted logistic regression analyses evaluating risk of poor outcomes (mRS of 3-6) at 90 days. (OR, (95% CI), P-value))

|  | All patients | Diabetic patients | Non-diabetic patients |
| --- | --- | --- | --- |
| Early Hyperglycemia | 1.90 (1.15-3.14),<br>**p=0.013** | 1.35 (0.39-4.72),<br>p=0.634 | 2.06 (1.17-3.63),<br>**p=0.012** |
| Late Hyperglycemia | 2.03 (1.14-3.62),<br>**p=0.017** | 2.44 (0.55-10.90),<br>p=0.244 | 1.86 (0.98-3.54),<br>p=0.057 |
| Persistent Hyperglycemia | 1.82 (1.32-2.52),<br>**p<0.001** | 1.45 (0.64-3.31),<br>p=0.371 | 2.14 (1.32-3.46),<br>**p=0.002** |

Reference group is persistent normoglycemia

**Table 3.** Adjusted logistic regression analyses evaluating risk of poor outcomes (mRS of 3-6) at 90 days. (OR, (95% CI), P-value))

|  | All patients | Diabetic patients | Non-diabetic patients |
| --- | --- | --- | --- |
| Early Hyperglycemia | 2.27 (1.10-4.68),<br>**p=0.026** | 3.63 (0.53-24.92),<br>p=0.189 | 2.86 (1.25-6.57),<br>**p=0.013** |
| Late Hyperglycemia | 1.64 (0.77-3.49),<br>p=0.200 | 4.39 (0.56-34.56),<br>p=0.159 | 1.60 (0.71-3.61),<br>p=0.261 |
| Persistent Hyperglycemia | 1.62 (0.95-2.79),<br>p=0.079 | 2.82 (0.74-10.82),<br>p=0.131 | 1.79 (0.94-3.43),<br>p=0.078 |

Reference group is persistent normoglycemia

### Subgroup analysis

In patients without diabetes, persistent normoglycemia had the highest proportion of patients with favorable functional outcomes at 46.4% (p<0.001). Late hyperglycemia group was also still associated with the lowest rate of survival probability at 90 days (85%) (p=0.005). In the unadjusted logistic regression, the association between poor functional outcomes with early hyperglycemia and persistent hyperglycemia groups remained significant (p=0.012 and p=0.003, respectively), while the late hyperglycemia group showed a non-significant trend (p=0.074). In the adjusted model, early hyperglycemia was still significantly associated with poor functional outcomes OR 2.86 (95% CI 1.25-6.57), p=0.013. In patients with diabetes, there was no statistically significant association between poor functional outcomes and any hyperglycemia group in either unadjusted or adjusted logistic regression. (Tables 2 and 3)

Upon stratifying our cohort into control and treatment arms, patients with persistent normoglycemia had the highest proportion of good functional outcomes in both subgroups at 45.7% and 45.9% (p=0.009 and p=0.030, respectively). Survival analysis revealed similar trends, with the late hyperglycemia patients showing the lowest chance of survival at 90 days in both the control arm and the treatment arm (p=0.007, p=0.402, respectively). In the univariable logistic regression model, all glycemic trajectories remained significant except early hyperglycemia trajectory in the control arm, and late hyperglycemia trajectory in the treatment arm p=0.323 and p=0.184, respectively. In the multivariable model, early hyperglycemia did not reach statistical significance in both the control arm and the treatment arm (p=0.184 and p=0.075, respectively).

## Discussion

In this secondary analysis of the ATACH-2 trial, we utilized serial measurements to characterize blood glucose trajectories during the first 72 hours after ICH. This method allows us to understand the fluctuations in glucose levels and assess their association with functional outcomes. Our findings demonstrate that distinct glucose trajectories were associated with divergent clinical outcomes. Early hyperglycemia independently predicted worse 90-day functional recovery, even after adjusting for ICH severity and relevant clinical variables. Late hyperglycemia was associated with the highest rates of hematoma expansion and the lowest probability of survival.

While prior studies have shown that hyperglycemia is associated with worse outcomes in ICH patients,^3,4,17,18,19,13^ conflicting data persists.^14,20^ Much of this inconsistency stems from a reliance on single glucose measurements, which fail to account for dynamic metabolic shifts that can occur after ICH. ^14,21,22,23^ Hyperglycemia occurs during the acute phase of ICH, when sympathetic activation and hypothalamic-pituitary-adrenal axis stimulation trigger massive catecholamine release, leading to insulin resistance and increased hepatic glucose output.^24^ Many unmeasured confounders, such as the timing and composition of the last pre-morbid meal, could influence the initial blood glucose measurement.^25^ By employing a trajectory-based model, we provide a more stable assessment of hyperglycemia as a marker of metabolic stress. Our results align with those of Wu et al., who identified that persistent hyperglycemia over 72 hours is a superior predictor of mortality compared to admission hyperglycemia alone.^21^

Preclinical models provide a biological support for the effect of hyperglycemia on brain injury. In animal studies, hyperglycemia has been shown to increase oxidative stress, facilitate the release of excitatory amino acids, and upregulate proinflammatory cytokines, including IL-1 and TNF-α. These processes promote microvascular injury and disrupt the blood-brain barrier, which leads to increased perihematomal edema and hematoma expansion.^26^ The inflammation can further worsen the effects of hyperglycemia in the brain and cause further metabolic dysregulation and secondary brain injury.^27,28^ Hyperglycemia can also cause direct damage to brain tissue by increasing lactate accumulation and intracellular acidosis, which could promote free radical formation and accelerate neuronal apoptosis. Zazulia et al. demonstrated that changes in perihematomal glucose metabolism peak at 72 hours,^29^ suggesting that metabolic response to injury and the window of vulnerability to hyperglycemia can last for several days. Our study found that the late hyperglycemia group had increased death. The mechanism of this is unclear, but it may be due to perihematomal growth, which has been associated with increased death.^30^

This further demonstrates the clinical relevance of the timing and duration of hyperglycemia as well as therapeutic window for intervention. While early hyperglycemia may reflect a catecholamine-mediated stress response,^22^ persistent and late hyperglycemia may indicate ongoing metabolic derangement.^21^

The prognostic significance of hyperglycemia may also differ depending on the pre-existing diagnosis of diabetes.^31^ Stress-induced hyperglycemia reflects an acute metabolic response in patients without diabetes and has been shown to be frequently associated with poor outcomes.^31,13^ The American Diabetes Association and the American Association of Clinical Endocrinologists, define stress hyperglycemia as any blood glucose level >140 mg/dL during a hospital stay in a patient without a history of diabetes.^13^ Stress hyperglycemia has been inconsistently demonstrated in diabetic patients ^3,22,32,13,5,33,34^ One reason for this inconsistency in findings may be the improved tolerance to hyperglycemia in diabetic patients due to their cellular adaptation to prolonged exposure to high blood glucose levels. In these patients, there is downregulation of GLUT-1 and GLUT-3 on endothelial cells and neurons, respectively, which limits the amount of glucose crossing the blood-brain barrier into the brain.^35^ Other reasons could include differences in confounders adjusted for in multivariable analysis, study populations, and unstressed baseline blood glucose levels.^31^ Diabetic patients may also be more likely to have better glycemic control.^20,36^ Nonetheless, our study found that hyperglycemia is significantly associated with poor outcomes in non-diabetic patients, but not in diabetic patients. This finding should be interpreted in the context of a relatively small diabetic sample size of only 18.6% of the study population.

Given our findings, it is important to review prior glucose-lowering trials. The NICE-SUGAR trial assessed blood glucose targets in critically ill patients and concluded that a target of 140-180 mg/dL resulted in lower mortality compared to 81-108 mg/dL.^37^ The GIST-UK trial assessed continuous infusion of glucose-potassium-insulin to achieve a blood glucose target of 72-126 mg/dL in stroke patients, but it did not show any benefit.^38^ The SHINE trial assessed intensive vs standard glucose control in ischemic stroke patients for 72 hours and determined that intensive control did not result in favorable functional outcomes at 90 days.^39^ However, the results of these trials are difficult to generalize to a population of patients with ICH, who are generally sicker.

Our study has several limitations. First, as a secondary analysis, the study was not primarily powered to detect differences in glucose trajectories, which may limit the statistical strength of certain analyses. Second, treatment protocols for hyperglycemia were not standardized across all ATACH-2 study centers, and we are uncertain whether patients who presented with hyperglycemia received treatment. Third, ATACH-2 trial excluded patients with low GCS scores, large hematomas, and those who did not undergo surgery; therefore, our results might be less generalizable to severe ICH. Fourth, baseline glycemic control was not available because our data does not include HbA1c, and some patients classified as stress hyperglycemia might have undiagnosed diabetes. Finally, blood glucose data were obtained intermittently at four time points over a 72-hour period, rather than continuously. As a result, we cannot account for glucose variability between these time points, which could influence mortality.^6^

## Conclusion

Results from our study demonstrate that the temporal pattern of hyperglycemia may predict prognosis following intracerebral hemorrhage. While early hyperglycemia likely reflects an acute sympathetic stress response, late and persistent hyperglycemia are more robustly associated with hematoma expansion and increased 90-day mortality. These findings suggest that glucose measurements at admission are insufficient for risk stratification and that clinicians should instead monitor blood glucose trajectories. Future prospective trials are also necessary to determine whether targeted interventions to maintain normoglycemia during the subacute phase of ICH (24-72 hours) can mitigate secondary brain injury and improve functional recovery in patients with ICH.

## Data Availability

ATACH-2 trial data was obtained from the NINDS biorepository

